# Health Communication Through News Media During the Early Stage of the COVID-19 Outbreak in China: A Digital Topic Modeling Approach

**DOI:** 10.1101/2020.03.29.20043547

**Authors:** Qian Liu, Zequan Zheng, Jiabin Zheng, Qiuyi Chen, Guan Liu, Sihan Chen, Bojia Chu, Hongyu Zhu, Babatunde Akinwunmi, Jian Huang, Casper J. P. Zhang, Wai-kit Ming

## Abstract

**Background:** In December 2019, some COVID-19 cases were first reported and soon the disease broke out. As this dreadful disease spreads rapidly, the mass media has been active in community education on COVID-19 by delivering health information about this novel coronavirus.

**Methods:** We adopted the Huike database to extract news articles about coronavirus from major press media, between January 1^st^, 2020, to February 20^th^, 2020. The data were sorted and analyzed by Python software and Python package Jieba. We sought a suitable topic number using the coherence number. We operated Latent Dirichlet Allocation (LDA) topic modeling with the suitable topic number and generated corresponding keywords and topic names. We divided these topics into different themes by plotting them into two-dimensional plane via multidimensional scaling.

**Findings:** After removing duplicates, 7791 relevant news reports were identified. We listed the number of articles published per day. According to the coherence value, we chose 20 as our number of topics and obtained their names and keywords. These topics were categorized into nine primary themes based on the topic visualization figure. The top three popular themes were prevention and control procedures, medical treatment and research, global/local social/economic influences, accounting for 32·6%, 16·6%, 11·8% of the collected reports respectively.

**Interpretation:** The Chinese mass media news reports lag behind the COVID-19 outbreak development. The major themes accounted for around half the content and tended to focus on the larger society than on individuals. The COVID-19 crisis has become a global issue, and society has also become concerned about donation and support as well as mental health. We recommend that future work should address the mass media’s actual impact on readers during the COVID-19 crisis through sentiment analysis of news data.

**Funding:** National Social Science Foundation of China (18CXW021)

**Evidence before this study:** The novel coronavirus related news reports have engaged public attention in China during the COVID-19 crisis. Topic modeling of these news articles can produce useful information about the significance of mass media for early health communication. We searched the Huike database, the most professional Chinese media content database, using the search term “coronavirus” for related news articles published from January 1st, 2020, to February 20th, 2020. We found that these articles can be classified into different themes according to their emphasis, however, we found no other studies apply topic modeling method to study them.

**Added value of this study:** To our knowledge, this study is the first to investigate the patterns of health communications through media and the role the media have played and are still playing in the light of the current COVID-19 crisis in China with topic modeling method. We compared the number of articles each day with the outbreak development and identified there’s a delay in reporting COVID-19 outbreak progression for Chinese mass media. We identify nine main themes for 7791 collected news reports and detail their emphasis respectively.

**Implications of all the available evidence:** Our results show that the mass media news reports play a significant role in health communication during the COVID-19 crisis, government can strengthen the report dynamics and enlarge the news coverage next time another disease strikes. Sentiment analysis of news data are needed to assess the actual effect of the news reports.

## Background

In December 2019, some pneumonia cases caused by an unknown pathogen were first reported in Wuhan, Hubei, China, and similar cases were soon reported in other provinces of China. The physicians and pathologists initially reported an “atypical pneumonia”, meaning a different type of pneumonia that was not caused by any of the three common pathogens, including *Streptococcus pneumonia, Hemophilus influenzae*, or *Moraxella catarrhalis*. After multiple sample collections and laboratory analyses, the pathogen was identified as a novel coronavirus, and the disease was named as COVID-19 by the World Health Organization (WHO) on February 11^th^, 2020 ^1^. The virus was named “SARS-CoV-2” by the International Committee of Taxonomy of Viruses ^2^.

Patients suffering from COVID-19 typically presented with fever, cough, and myalgia or fatigue. Less common manifestations included sputum production, headache, hemoptysis, and diarrhea. Abnormal chest CT images were found in all patients, and more than 50% of patients developed dyspnea. Some patients in critical condition might even die ^3^. Evidence shows that droplet transmission through the respiratory tract and close contact transmission are the two main transmission routes of COVID-19, while the aerosol transmission is a possible transmission route when people are exposed to high concentrations of virus aerosol in a relatively closed environment for some time ^4^. According to the National Health Commission (NHC) of the People’s Republic of China, until February 2020, there had been approximately 80000 confirmed cases and more than 2000 deaths in China ^5^. Other countries, such as Japan, South Korea, Thailand, Singapore, and the United States, also reported COVID-19 cases in their countries ^6^. Although the cases at the early stage in these countries were identified as imported cases from Wuhan or other cities in Hubei province, some domestic cases and local transmission have also been reported.

The rapid spread of COVID-19 has already caused great public attention, and many heated discussions, and the Chinese mass media have been reporting relevant information about the virus and the outbreak. As effective public health measures are required to be implemented in time to avoid the breakdown of the health system ^7^, the media certainly can play a crucial role in conveying updated policies and regulations from authorities to the citizens.

Besides, since no COVID-19 vaccine is yet available, each citizen should be aware of the harm caused by this novel coronavirus, the prevention methods, and the designated hospital in their local area to access at any point in time. If misleading or incorrect information was transmitted to the public, citizens might feel panic and thus make a panic purchase and try unnecessary or even detrimental medicine regimens. Therefore, it requires the mass media information dissemination activities in conjunction with the health stakeholders to help individuals, authorities, government, and other stakeholders to understand the precarious global and public health condition posed by the COVID-19 and identify health-related knowledge and training required in facing the menace.

Given the desire to know whether the media work efficiently in delivering the latest COVID-19 information to the public audience, Major media reports were collected and analyzed. Multimodal data modeling can combine multiple information from various resources. To cope with multimodal data, topic modeling is used. Topic modeling is a type of statistical model that arranges unstructured data structurally in accordance with latent themes. In this way, we could investigate the patterns of health communication through the media and the role the media that have played so far during the COVID-19 crisis in China.

## Methods

### Data collection

We collected Chinese news and related articles on COVID-19 from January 1^st^, 2020, to February 20^th^, 2020. We then applied the Latent Dirichlet Allocation (LDA) modeling method to derive useful information from these news reports.

Chinese news and related articles data were collected from the Huike database. The Huike database is one of the most professional, ever-growing Chinese media content databases, containing the news and article data from more than 1500 print media and over 10,000 internet media. The news and articles data in the Huike database are updated in a timely manner ^8^.

To gain insights into the early period of health communication information about coronavirus, we conducted a search with the keyword “coronavirus” in the Huike database.

LDA is a generative probabilistic topic modeling method that is widely applied in text mining ^9^, medicine ^10,11^, and social network analysis ^12^ due to its excellent capability of converting visual words into images and visual word document ^13-15^. It is a generative statistical model with a three-level hierarchical Bayesian model. The basic assumption of this model is a combination of words belonging to different topics ^16^. LDA indicated that there may be various topics in an article and that the wording in that article is attributable to one of its topics. We can discover the topics among the data pool by using Gibbs Sampling techniques ^17^.

### Processing

Eleven-thousand-two-hundred-twenty articles were found with keyword search “coronavirus”, dated between January 1^st^, 2020 and February 20^th^, 2020. After cleaning the data, 7791 articles remained.

Before applying LDA modeling, we used Python to perform data cleaning and used the Python package Jieba for data process ^18,19^. The detailed data process is illustrated in Figure 1. We next removed Chinese common stop characters, such as “ten”, “a”, “of”, and “it”. We also built a document-term matrix (DTM) and used TF-IDF to process the data.

**Figure 1:**
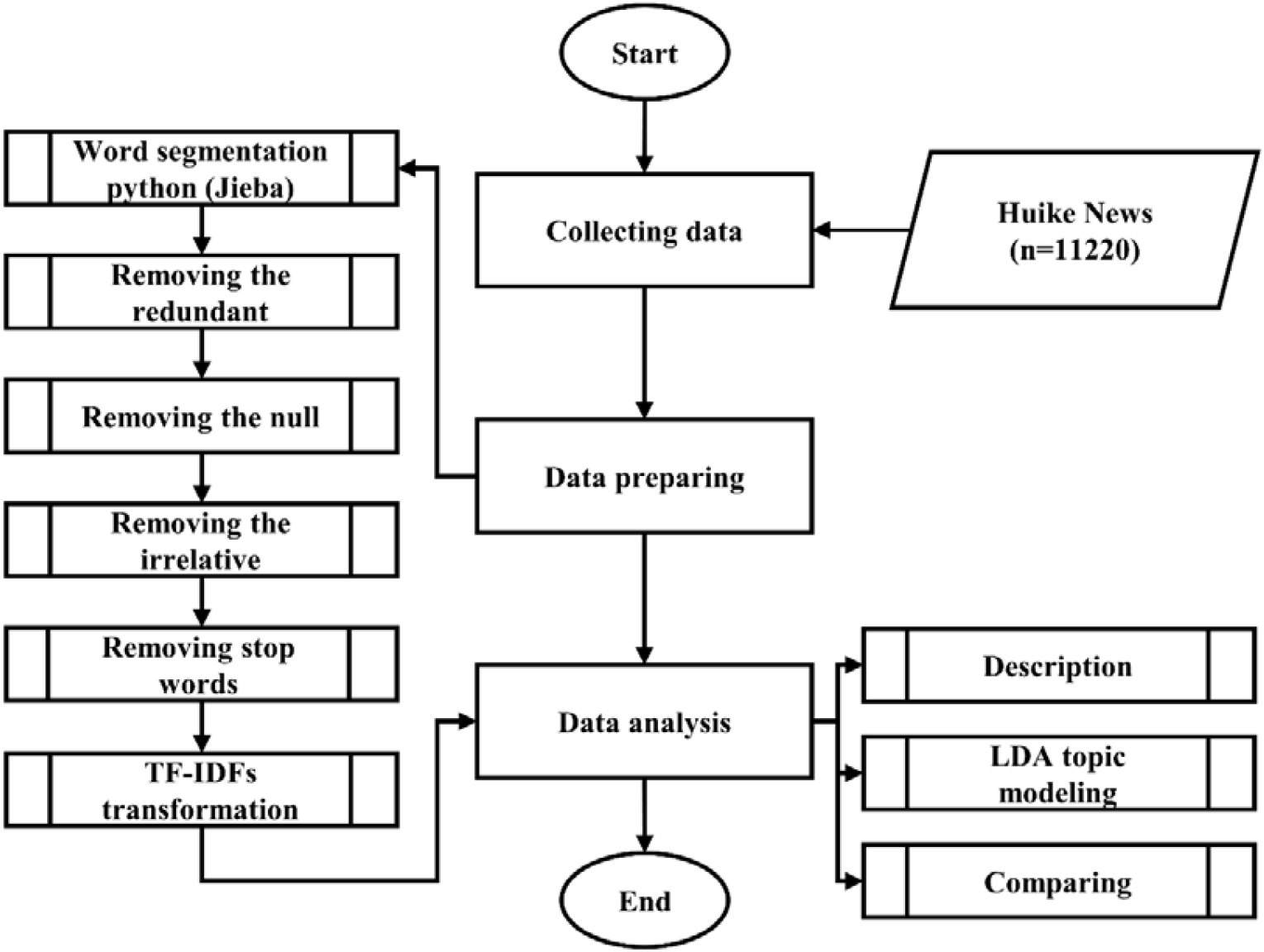
Data processing flow chart.

To seek a suitable LDA topic number and the explanations to investigate the relationship between COVID-19 crisis and news reports, we conducted multiple studies. For the selection of a suitable number of topics, we used a coherence score to evaluate ^20^. Topic coherence measures the consistency of a single topic by measuring the semantic similarity between words with high scores in a topic, which contributes to improving the semantic understanding of the topic. That is, words are represented as vectors by the word co-occurrence relation, and semantic similarity is the cosine similarity between word vectors. The coherence is the arithmetic mean of these similarities ^21^. We used Coherence Model from Gensim, the python package for natural language processing, to calculate coherence value ^22^. According to Figure 2, the coherence score increased and reached a stable score as the number of topics increased to 20, then declined after the number of topics reached 25. However, we found that the result could be uninterpretable for humans if only statistical measures were applied ^23^. As a result, we combined statistical measures and manual interpretation and chose 20 topics to analyze with the help of Python 3.6.1 version and LDAvis tool ^16^. We set λ = 1 and set 20 topics and their keywords. Topics’ names were generated according to their corresponding keywords to expatiate the topics.

**Figure 2:**
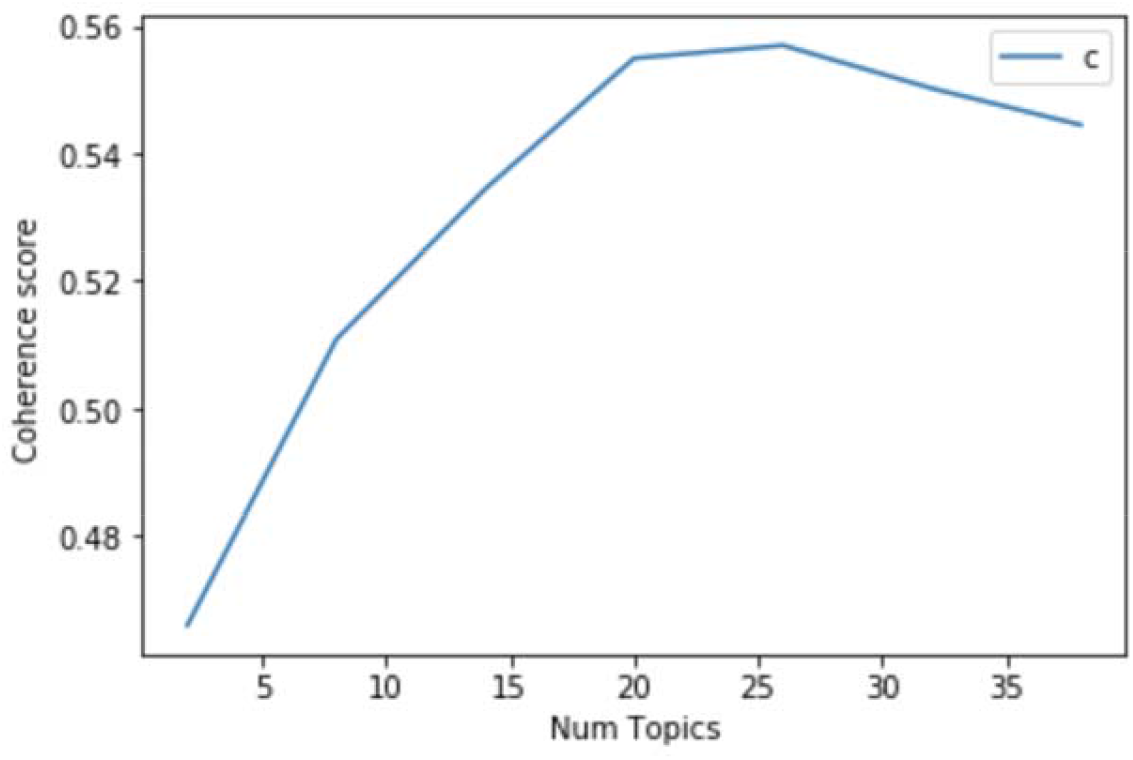
Coherence score for the topic numbers.

We also divided these topics into different themes to study them better. In the visualization, as in the two-dimensional plane (Figures 3 and 4), 20 topics were represented as cycles. These circles overlapped, and their centers are determined by computed topic distance ^16^. By this approach, these 20 topics were classified into nine main primary themes and are shown in Table 1.

**Table 1:** Topic classification and keywords.

| Theme | Topic names | Keywords | Topic proportion | Theme proportion |
| --- | --- | --- | --- | --- |
| Theme 1: Confirmed cases | Topic 5 | Cases, Confirmed, Patient, Pneumonia, Novel, Coronavirus, Infection | 5·7 | 9·6 |
| .. | Topic 17 | New type, Novel coronavirus, Pneumonia, Infection | 3·9 | .. |
| Theme 2: Medical supplies | Topic 7 | Mask, Disinfection, Protection, Contact, Symptom | 5·6 | 5·6 |
| Theme 3: Medical treatment and research | Topic 16 | Detection, Research, Laboratory, Treatment, Coronavirus, Drug | 4·2 | 16·1 |
| .. | Topic 4 | Virus, Infection, Spread | 6·4 | .. |
| .. | Topic 8 | Hospital, Patient, Medical staff, Wuhan, Medical team | 5·5 | .. |
| Theme 4: Prevention and control procedures | Topic 1 | Prevention and Control, Work, Meeting, Outbreak, Fight against the outbreak, Conference | 7·2 | 32·6 |
| .. | Topic 6 | Personnel, Prevention and Control, Community, Outbreak, Quarantine | 5·6 | .. |
| .. | Topic 10 | Prevention and Control, Work regulation, Department, outbreak, Measure in accordance with the law, Quarantine | 4·8 | .. |
| .. | Topic 3 | Prevention and control, Measures, Outbreak | 6·5 | .. |
| .. | Topic 19 | Outbreak, Company, Prevention and control, Coronavirus, Pneumonia, Impact, Fight against, Employee | 3·7 | .. |
| .. | Topic 9 | Enterprise, Outbreak, Service, Prevention and Control, Guarantee, Support, Manufacture | 4·8 | .. |
| Theme 5: Wuhan's story | Topic 2 | Wuhan, Work, Spring festival, Front line, Family member, Together | 6·7 | 6·7 |
| Theme 6: Mental issue | Topic 14 | Outbreak, Information, Mental, Society, Outbreak, Platform, People, Nationwide, Epidemic control | 4·4 | 4·4 |
| Theme 7: Global / local social / economic influences | Topic 20 | Hong Kong, Mainland, Taiwan, Macao, Pneumonia, outbreak, Government, Impact | 3·7 | 11·8 |
| .. | Topic 18 | Cancel, Event, Hotel, Visitor, Spring festival, Tourism, Announce, Journalist, Wuhan | 3·8 | .. |
| .. | Topic 15 | China, International, Response, Take measures, Outbreak | 4·3 | .. |
| Theme 8: Materials supplies and society support | Topic 13 | Materials, Donation, Mask, Wuhan, Anti-attack, Medical, Prevention and Control, Hubei | 4·4 | 8·9 |
| .. | Topic 11 | Mask, Production, Enterprise, Supply, Price, Manufacture, Market | 4·5 | .. |
| Theme 9: Detection at public transportation | Topic 12 | Passenger, Wuhan, Body temperature, Detection, Airport, Vehicle | 4·4 | 4·4 |

Remarks: The total percentage is 100·1% due to automatic rounding while exporting the results.

**Figure 3:**
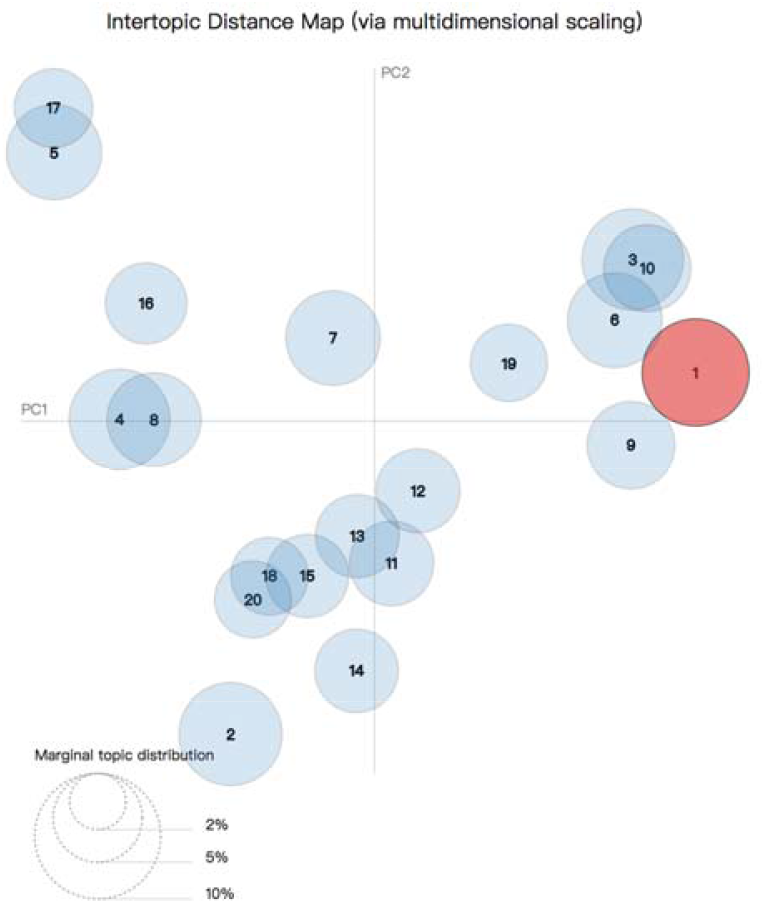
Inter-topic distance map.

**Figure 4:**
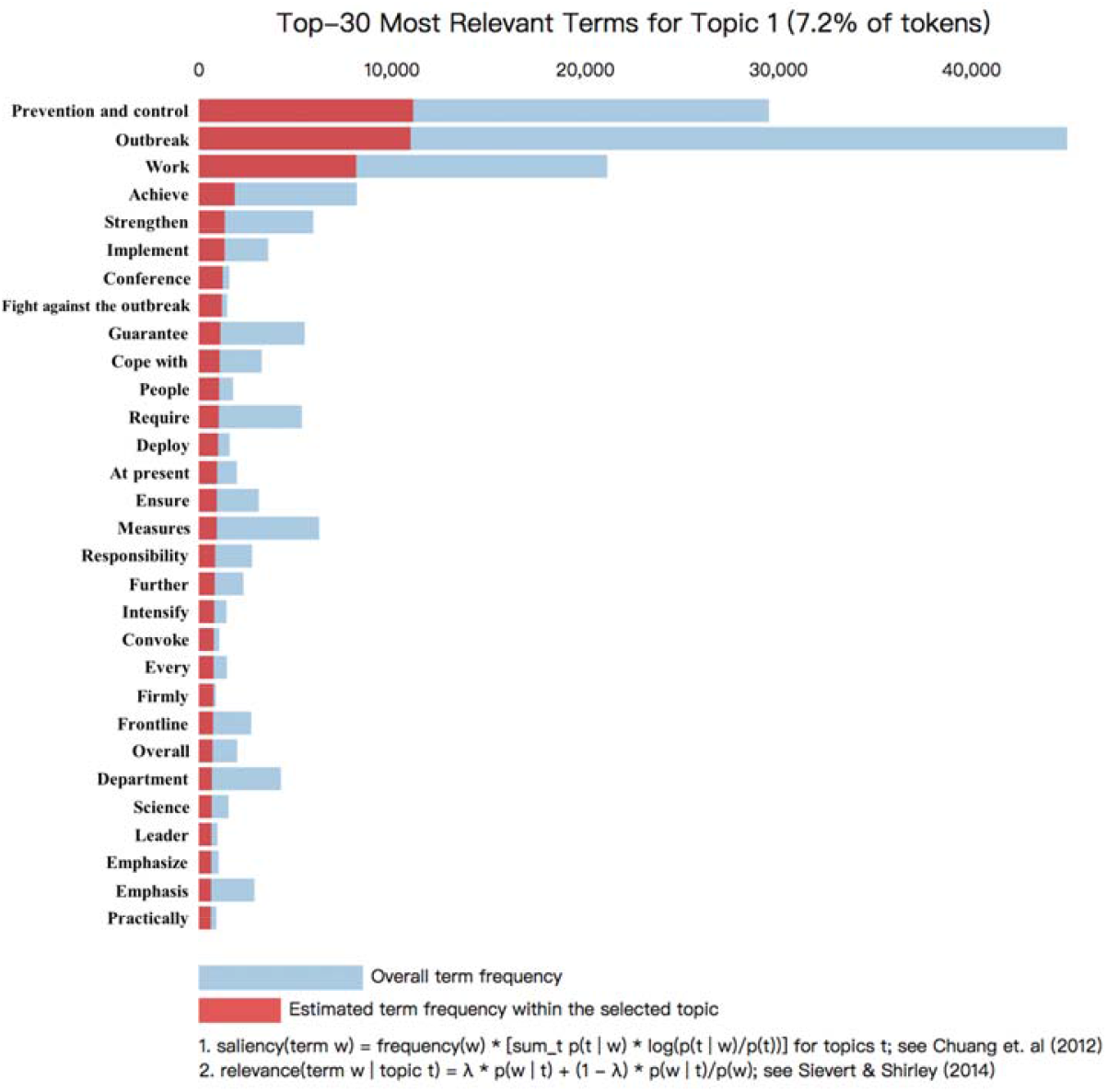
Top-30 most relevant terms for Topic 1.

## Results

Figure 3 shows the design of the topic model, in which 20 different topics are plotted as circles. The areas of the circles indicate the overall prevalence, and the center of the circles is determined by computing the distance between topics. Intertopic distances are shown on a two-dimensional plane ^24^ via multidimensional scaling. PC1 represents the transverse axis, and the PC2 represents the longitudinal axis.

Figure 4 shows the top 30 most relevant terms for interpreting Topic 1. Each bar shows the given term’s overall frequency and the estimated frequency within Topic 1. This approach is illustrated in the literature ^25^.

Figure 5 shows that the number of relevant news slightly increased after a new death was reported on 9 January 2020. Between 20 to 23 Jan 2020, we observed a sharp increase of relevant news. As the daily news cases decreased since 4 Jan 2020, the number of daily news reports began to drop. The increase in the number of cases on 12 and 13 Feb was due to the updated diagnosis criteria in the COVID-19 protocol (fifth version) ^26^.

**Figure 5:**
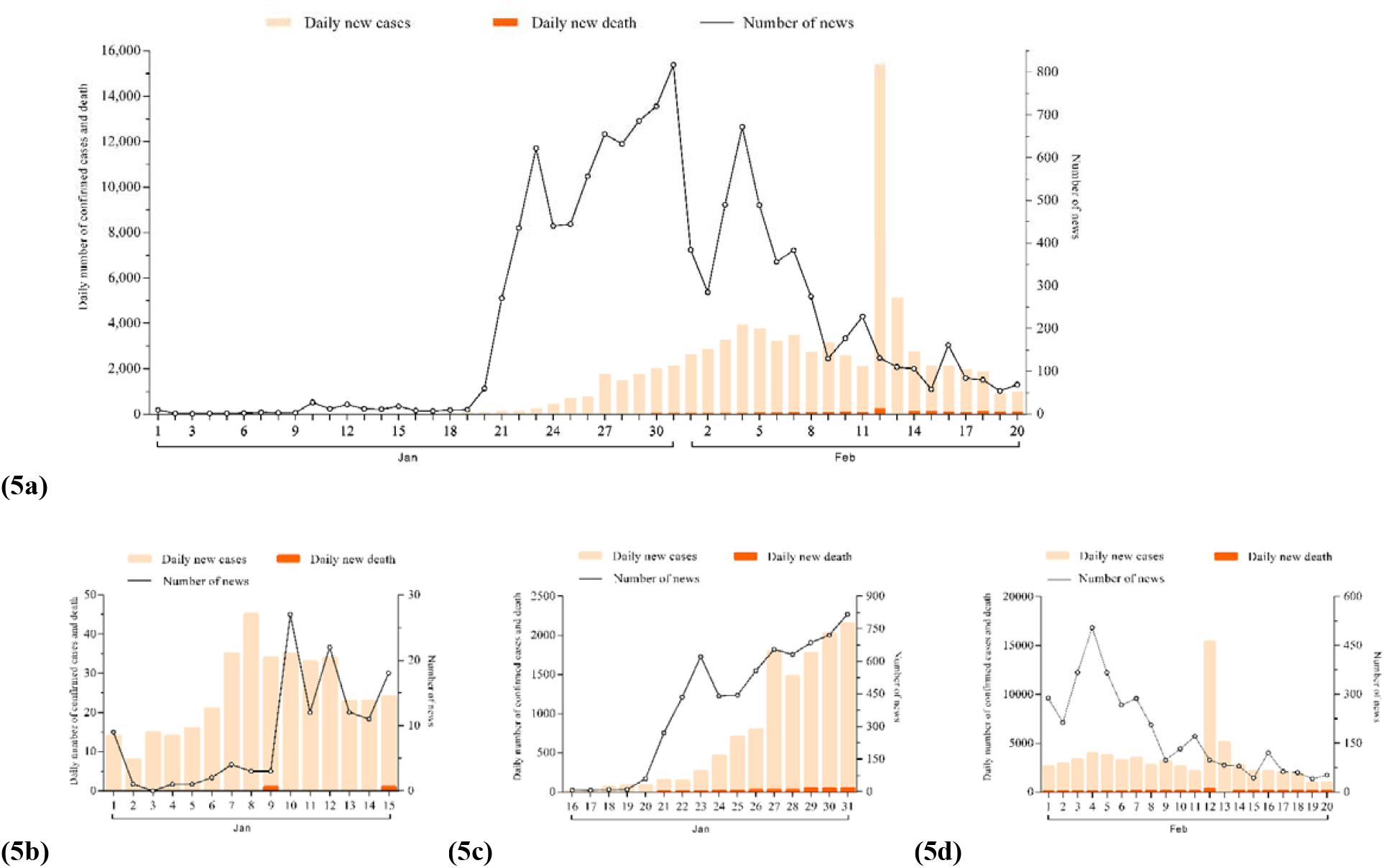
Time series news streams with daily confirmed cases and death. Remarks: Figures 5b, 5c and 5d are the partial magnification of the Figure 5a. The data of daily confirmed cases and death between January 1^st^, 2020, to January 16^th^ was extracted from the figure in a transmission dynamics study published at March 26^th^, 2020 ^27^.

**Figure 6:**
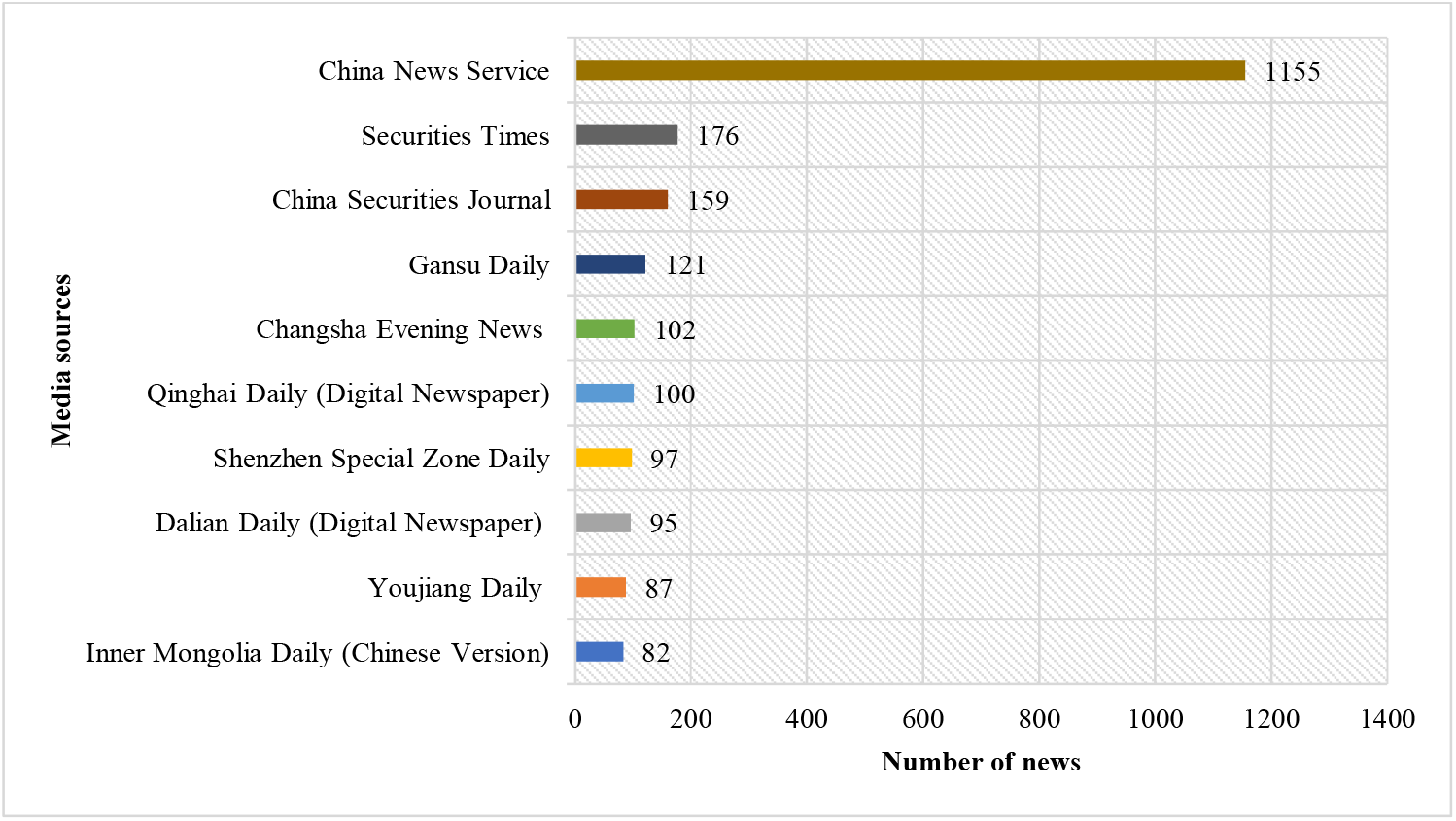
The most represented media sources. As demonstrated in Figure 6, China News Service was the most productive media source, followed by the Securities Times and China Securities Journal. Local and national newspapers all participated in reporting recent updates.

**Figure 7:**
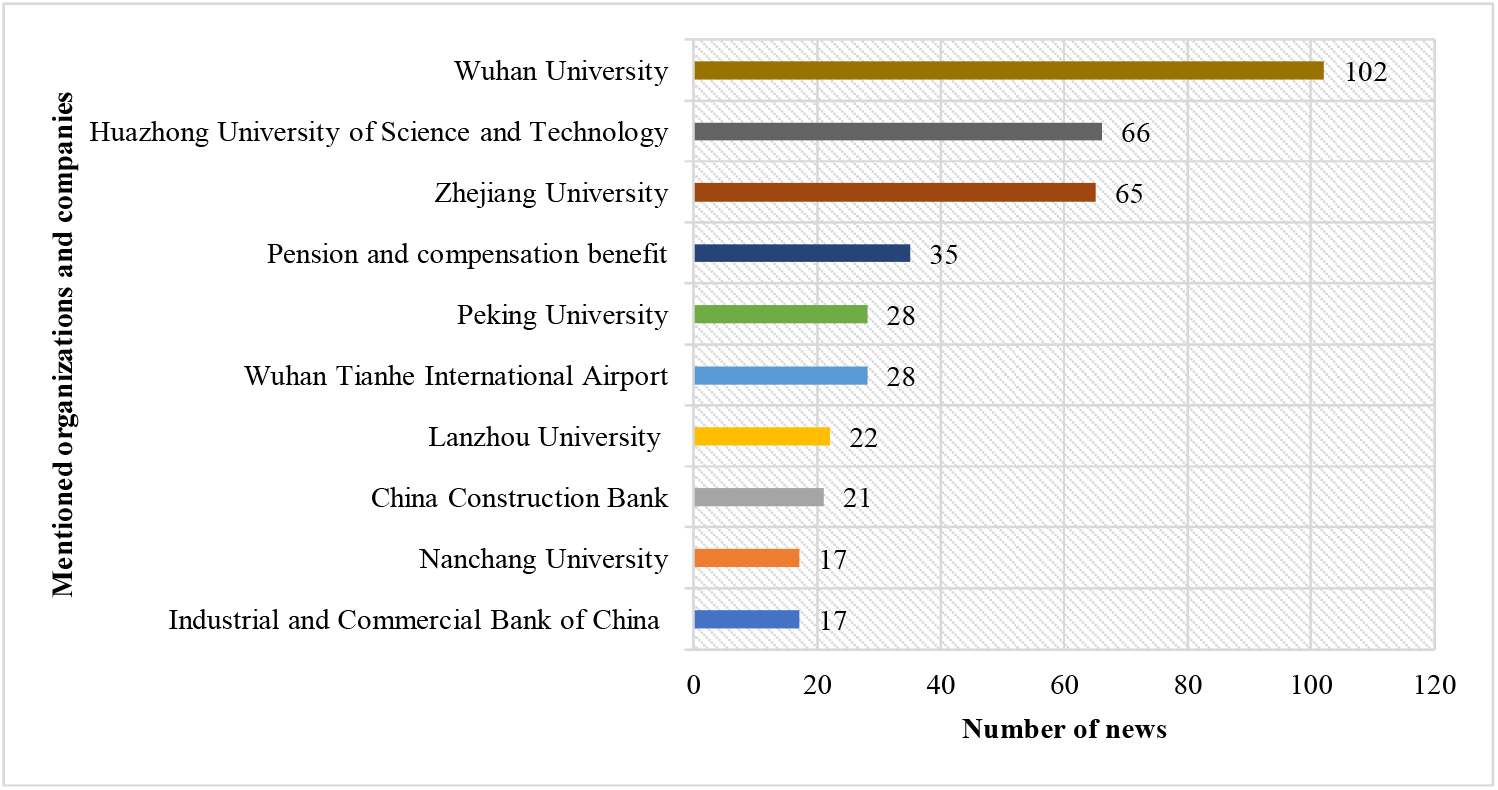
Organizations and companies mentioned in news reports. Our collected news reports mentioned various organizations and companies as shown in Figure 7. Wuhan University and Huangzhong University of Science and Technology, as two top universities in Wuhan, have been the most mentioned, followed by Zhejiang University. University affinitive hospitals and University alumni association participate actively in the fight against COVID-19.

Figure 8 shows the theme percentage allocation of our collected news reports. Given our analysis, Theme 4 (Prevention and control procedures) was the most popular theme, accounting for 32·6% of the total. Theme 3 (Medical treatment and research) was involved in about 16·1% of the related news. Theme 7 (Global/local social/economic influences) was included in 11·8% of all news reports of coronavirus. The other six themes each account for less than 10% of news stories. Theme 1 (Confirmed cases) was related to 9·6%. Themes 8 (Materials supplies and society support), 5 (Wuhan story), 2 (Medical supplies), 6 (Mental issue), and 9 (Detection at public transportation) accounted for 8·9%, 6·7%, 5·6%, 4·4%, and 4·4% of articles, respectively.

**Figure 8:**
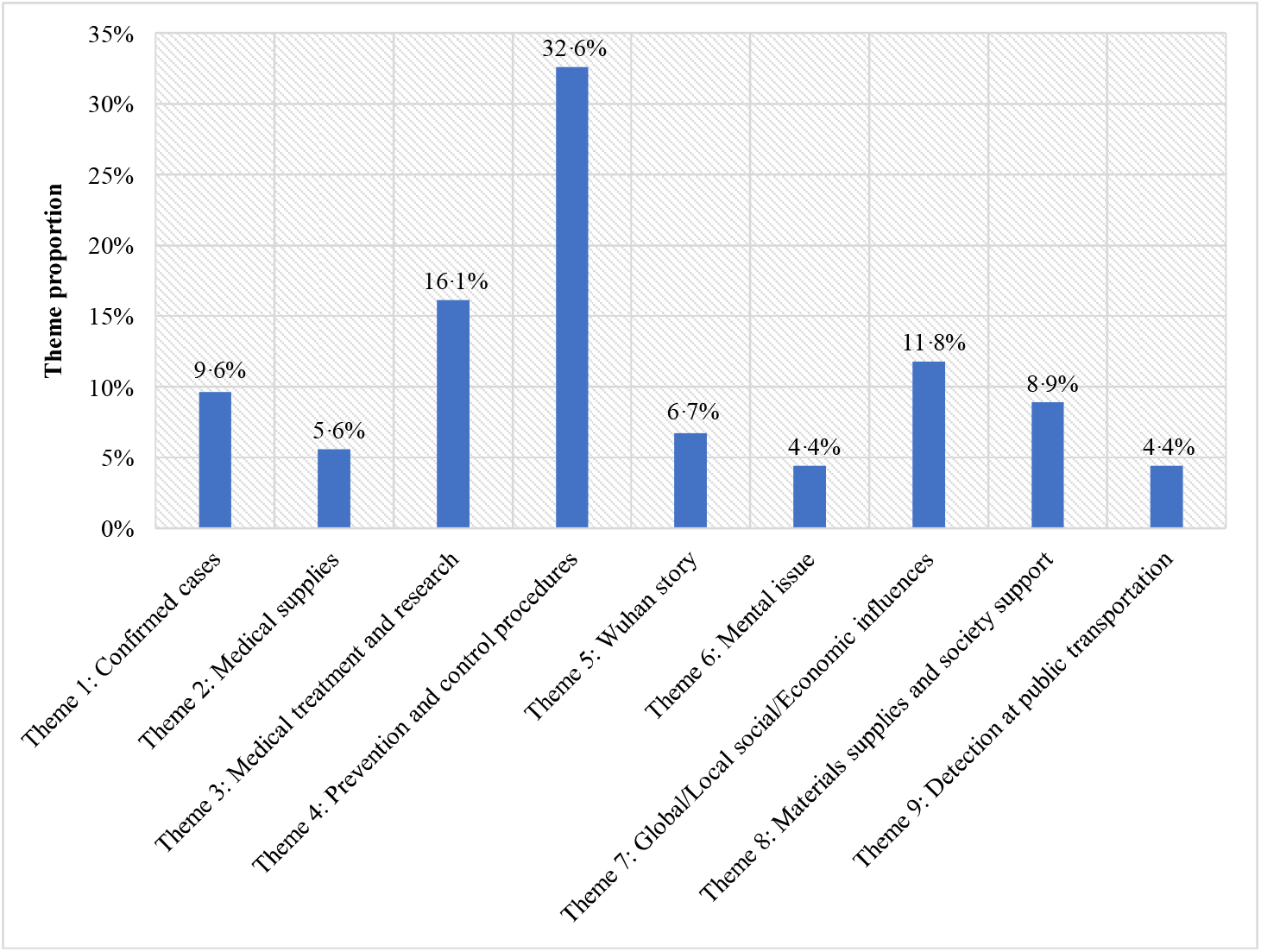
Theme percentage allocation. Remarks: The total percentage is 100·1% due to automatic rounding while exporting the results.

## Discussion

The COVID-19 crisis has aroused great public concern not just in China but also around the world. Topic modeling provides an alternative perspective to investigate the relationship between media reports and the COVID-19 outbreak. We collected media reports and used topic modeling to analyze them. Although several COVID-19 cases were found in December 2019, we observed few news reports about it, showing that the press media did not focus on this disease at that time. As the outbreak became severe and more pneumonia cases were confirmed, the number of news reports began to steadily increase, then rapidly increase on January 19^th^, 2020. It takes time and rigorous effort for journalists to choose a topic, investigate the situation, collect data, and verify the authenticity of the material before they can finally present the news. As a result, a delay is unavoidable in most cases. Therefore, mass media news reports lag behind the real-time coronavirus developments, indicating that the media does not play an adequately forewarning function in public health communication and sensitization.

The topics focused by mass media can be divided into nine classifications. Theme 4 (Prevention and control procedure), and Theme 3 (Medical treatment and research) are two major themes that together accounted for around half of the content. The management of important government departments and medical institutions and community control methods are also emphasized. Positive and enthusiastic forecasts backed with active public health interventions are disseminated, which can eliminate the unnecessary public worry and extreme panic, in the meantime aimed at asserting the nation’s confidence in virus containment and victory within a short period.

Control of sources of infection, interruption of transmission routes, and protection of susceptible people are three major principles to prevent and control infectious diseases. To cope with the COVID-19 crisis, the Chinese government also take measures based on these three principles. The detection of viral infections within public transportation networks aroused great public concerns, given that the outbreak coincided with the spring festival, when many are traveling. Few news reports about this are included in Theme 4 (Prevention and control procedure), suggesting that the mass media might not have been providing sufficient health information about detection within the transportation network.

The scale of medical treatments and research was the second most popular topic. Our results show that the mass media convey this kind of health information by paying attention to the detection of suspicious cases, drugs that might cure patients, and the transmission route of the virus. However, reports about Theme 4 (Prevention and control procedure) and Theme 3 (Medical treatment and research) mainly focus on the whole society level, while instructions on personal prevention and clinic and medicine choices are less mentioned.

Influences on activities (home and abroad) were also reported, together with economic influences, included in Theme 7 (Global/local social/economic influences). These data indicate that the impact of the COVID-19 crisis is not limited to the medical field, but also social and economic fields. It is also a global health issue that requires people around the world to work closely together.

The term “confirmed cases” appeared in 9·6% of the articles. This indicates that the mass media has served a public health function because case number and its changing rates in news reports can directly give the public intuitive feelings about the speed of viral spread, momentum, and the hazard of this coronavirus. It can also help citizens remain alert about virus transmission and, therefore, change their daily habits accordingly.

Theme 2 (Medical supplies) and Theme 8 (Materials supplies and society support) connect the material supplies with the COVID-19 crisis. Since the outbreak is so sudden and the transmission is so rapid, people in affected areas require medical material and other necessities, especially after the Chinese government shut the major entrance to Hubei to control the outbreak. The mass media can communicate with other parts of China to call on donations and support.

There is an emphasis on Wuhan’s story, where news stories focus on an individual’s life instead of the whole city level. We also observed that Theme 6 (Mental health) accounts for 4·4% of all news articles, revealing that there is a need for mental health intervention of patients, medical staff, and the public in the news reports. These two themes indicate the mass media adopts a people-oriented principle when reporting the COVID-19 crisis.

### Limitations

In our study, we included a large number of Chinese news articles about COVID-19 from the Huike mass media database which only covers text news articles. However, the mass media recently used new media platforms such as Tik Tok and WeChat (the largest Chinese instant messaging application) to deliver health information through images, snapshots, and short videos. Therefore, we might omit the news content and the impact of mass media on these media platforms. Besides, our study mainly researches the mass media’s role; it would be valuable if we could measure the public’s reaction to our study.

## Conclusion

Collecting and analyzing reports on the novel coronavirus shed light upon how the Chinese media have delivered health information during the COVID-19 crisis. Our study provides evidence that the Chinese mass media news lags behind when reporting the major developments of the viral spread. Prevention and control procedures, medical treatment, and research are major themes of the press, but mainly focusing on the whole society level, while instructions on personal and individual prevention, clinic and medicine choices, and detection need to be further enhanced. Global and local influences had been reported as the COVID-19 crisis started to impose pressure on public health globally and urge cooperation among all humankind. Further research should be considered to explore the impacts of mass media on the readers through sentiment analysis of news data and the influences of misinformation about COVID-19 delivered through the mass media.

## Contributors

Qian Liu and Wai-kit Ming conceived the original idea, designed the whole research process.

Qian Liu, Quan Liu, Qiuyi Chen collected and cleaned data

Qian Liu and Wai-kit Ming did the data analysis, data interpretation, and wrote the first version of the manuscript.

Qian Liu, Jiabin Zheng, Zequan Zheng made the figures.

Sihan Chen, Bojia Chu, Hongyu Zhu, Jian Huang, Casper J. P. Zhang, Babatunde Akinwunmi contributed to the administration of the project and data analysis, data interpretation.

Both Zequan Zheng and Jiabin Zheng contributed to the final version of the manuscript.

Babatunde Akinwunmi and Wai-kit Ming reviewed the manuscript.

All authors contributed to the interpretation of the results and final manuscript.

All authors discussed and agreed on the implications of the study findings and approved the final version to be published.

## Data Availability

Data are available from existing online repositories that are listed in the manuscript.

